# Tau Burden is Best Captured by Magnitude and Extent: Tau-MaX as a Measure of Global Tau

**DOI:** 10.1101/2025.01.13.25320488

**Authors:** Christopher A. Brown, Sandhitsu R. Das, Katheryn A.Q. Cousins, Thomas F. Tropea, Alice-Chen Plotkin, John A. Detre, Paul A. Yushkevich, Corey T. McMillan, Edward B. Lee, Leslie M. Shaw, Ilya M. Nasrallah, the Alzheimer’s Disease Neuroimaging Initiative, David A. Wolk

## Abstract

Tau exhibits change in both spatial extent and density of pathology along the Alzheimer’s disease (AD) spectrum with each aspect contributing to the overall burden of pathological tau. Nevertheless, studies using Tau PET have measured either magnitude using standardized uptake value ratios (SUVRs) or extent using number of Tau+ regions. We hypothesized that combining these two dimensions into a single measure of Magnitude and eXtent, Tau-MaX, would provide improved quantification of global tau burden as well as allowing for a region-agnostic measure of global tau burden that does not require a pre-specified region of interest (ROI) or meta-ROI. To test this hypothesis, we analyzed ^18^F-flortaucipir PET scans from local and national consortium data (n=1077 participants total) and used Gaussian-mixture models for data from 64 brain regions, to define both tau positivity and magnitude. We examined cross-sectional and longitudinal change in Tau-MaX across the Alzheimer’s disease (AD) spectrum and compared the association of Tau-MaX, magnitude, and extent with plasma p-tau_217_ and global cognition. We also compared Tau-MaX using a global, region-agnostic approach to temporal lobe or Braak stage meta-ROIs. Whereas separate assessments of extent and magnitude across the disease spectrum found earlier increases in Tau spatial extent and later increases in magnitude, Tau-MaX was able to dynamically capture this shift demonstrating a stronger association with extent in the preclinical stage and a stronger association with magnitude in clinical stages. Global Tau-MaX differed between disease stages cross-sectionally and changed over time in all stages of disease. Further, Tau-MaX significantly improved associations with plasma p-tau_217_ and global cognition compared to magnitude or extent alone. Finally, global measures of Tau-MaX performed similarly to meta-ROI measures of Tau-MaX. Together, these findings indicate that combining magnitude and extent provides a robust measure of global tau burden that changes throughout the disease course and is associated with blood-based biomarkers and cognition. This measure may be of particular use for disease staging, as well as serving as an outcome measure to monitor response to therapeutic intervention.

## Introduction

Over the last decade, the availability of tau PET tracers has enabled *in vivo* quantification of tau pathology across the Alzheimer’s disease (AD) spectrum. Initial studies focused on the ability to recapitulate the autopsy-defined Braak staging pattern, starting with binding in transentorhinal and hippocampal regions of the medial temporal lobe (MTL), followed by spread to neocortical temporal regions and later to parietal, frontal, and occipital cortices^1–4^. Additional studies have also examined deviations from this canonical pattern of tau spread to identify specific sub-types that closely resemble those described at autopsy, as well as heterogeneity within canonical and atypical subtypes^5–7^. While there has been great interest in exploring this heterogeneity in regional tau burden, particularly focusing on propagation within brain networks^8–10^, methods of staging disease based on tau burden have largely adhered to a homogenous approach with pre-specified regions in the temporal lobe as early sites of tau deposition and neocortical parietal, frontal, and occipital regions used as a measure of later stage disease^11–13^.

The most common approach involves using standardized uptake value ratio (SUVR) in a temporal meta-ROI, consisting of MTL and a subset of temporal neocortical regions, as a measure of tau severity in early disease, and average SUVR across a set of Braak stage 4-6 regions to assess tau severity in moderate and late disease stages^11,14^. While this approach may correctly quantify tau severity in the majority of individuals who follow canonical patterns, it may still inaccurately assess the burden in those who deviate from this pattern. One solution to this problem of heterogeneity is to assess the degree of tau positivity across all brain regions within an individual without applying a pre-conceived pattern of regional involvement, allowing for calculation of a Spatial Extent Index^15^. Recent work using this approach in the Alzheimer’s Disease Neuroimaging Initiative (ADNI), found that this region-agnostic approach performs similarly to temporal meta-ROI SUVR for predicting memory performance and outperforms it in predicting executive function performance^15^.

While the Spatial Extent Index demonstrates the value of using a region-agnostic approach to consider spatial extent of tau pathology, it does not account for differences in magnitude of tau burden across regions and manifests ceiling effects in individuals with dementia^15^. Furthermore, while spatial extent is akin to the Braak staging used in postmortem studies, this approach has limitations for disease staging due to the consistently reported large variability in tau tangle density across and within Braak stages^16–18^. For example, later Braak stages may be associated with less tau load in earlier Braak regions than cases who are in earlier stages.

Extending the Spatial Extent Index to incorporate magnitude of tau burden within tau positive regions might be expected to provide an improved measure of overall tau burden. To test this hypothesis, we used tau PET scans from the Alzheimer’s Disease Neuroimaging Initiative (ADNI, n = 892) and our University of Pennsylvania Alzheimer’s Disease Research Center (Penn ADRC, n = 180) Aging Brain Cohort (ABC) to characterize spatial extent and magnitude of tau pathology. We employed Gaussian-mixture models to calculate Tau Pathology Index (TPI) as a measure of magnitude and cut-points of two standard deviations above the mean of the non-pathological curve as a measure of tau positivity. We combined these two dimensions by adding the TPIs of all tau+ regions to form a new measure of Tau Magnitude and eXtent (Tau-MaX). We assessed Tau-MaX cross-sectionally and longitudinally across the AD spectrum, and we correlated Tau-MaX with plasma p-tau_217_ and global cognitive measures. We compared Tau-MaX to SUVR and extent alone using a fully region-agnostic global approach, as well as within temporal and Braak stage meta-ROIs.

## Materials and methods

### Participants

We used tau PET from the Penn ABC and ADNI cohorts and included all individuals with at least one ^18^F-flortaucipir PET scan and MRI prior to July 2024 (ADNI) or March 2024 (ABC). A subset of participants had available Amyloid PET using either ^18^F-Florbetaben or ^18^F-Florbetapir, Mini-mental Status Examination (MMSE) or derived MMSE from the Montreal Cognitive Assessment (MoCA), and/or Fujirebio Lumipulse plasma p-tau_217_. A subset of participants from ADNI also had neuropsychological composite test scores for memory, language, executive function, and visuospatial function. All participants in ADNI and ABC underwent study-specific consensus diagnosis to determine their clinical status as: 1) Cognitively Unimpaired (CU), 2) Mild Cognitive Impairment (MCI), 3) Dementia, or 4) Impaired-Not MCI. In accordance with the Declaration of Helsinki, all participants provided informed consent under protocols approved by local Institutional Review Boards. Details about ADNI informed consent procedures are available from www.adni-info.org.

A subset of data used in the preparation of this article were obtained from the Alzheimer’s Disease Neuroimaging Initiative (ADNI) database (adni.loni.usc.edu). The ADNI was launched in 2004 as a public-private partnership, led by Principal Investigator Michael W. Weiner, MD. The primary goal of ADNI has been to test whether serial magnetic resonance imaging (MRI), positron emission tomography (PET), other biological markers, and clinical and neuropsychological assessment can be combined to measure the progression of mild cognitive impairment (MCI) and early Alzheimer’s disease (AD). For up-to-date information, see www.adni-info.org.

### PET Acquisition and Processing

For both ADNI and ABC, tau PET was acquired using six 5-minute frames from 75-105 minutes after injection of ^18^F-Flortaucipir. Processed ^18^F-Flortaucipir PET images with uniform 6mm full-width-at-half-maximum resolution were downloaded from the ADNI archive (“Coreg, Avg, Std Img and Vox Size, Uniform Resolution”). An in-house processing pipeline was implemented to replicate these processing steps for ABC data with registration of all frames to the first image, averaging, resampling to 1.5mm^3^ AC-PC aligned space, and smoothing with uniform 6mm full-width-half-maximum resolution. All PET data were registered to T1-weighted images using Advanced Normalization Tools (ANTs) rigid-body registration. All registrations were visually checked for quality control using standardized slices and grids. T1-weighted images were bias-corrected and skull-stripped using ANTs prior to cerebellar, cortical, and subcortical parcellation using multi-atlas segmentation with Joint Label fusion with the MICCAI 2012 BrainColor parcellation, which generates 102 cortical ROIs^19–22^. An inferior cerebellar reference region was used to generate SUVR maps and mean SUVR was extracted from all cortical/subcortical regions after partial volume correction (excluding other cerebellar and brainstem regions).

For both ADNI and ABC, amyloid PET was acquired using four 5-minute frames from 90-110 minutes after injection of ^18^F-florbetaben or 50-70 minutes after injection of ^18^F-florbetapir. Amyloid status (Aß+/-) was assessed by visual read by a trained nuclear medicine physician (I.M.N.) for ABC. For ADNI, SUVR in a neocortical composite region derived by the ADNI PET core was used to determine Aβ positivity with the recommended cutoff value of > 1.11 SUVR for florbetapir or > 1.08 for florbetaben.

### Regional Tau PET Measures

We used Gaussian mixture models to differentiate off-target from on-target binding^8,9^. Regions were selected by excluding region-pairs better fit by a single-Gaussian model in either hemisphere. For the remaining 64 regions (covering 73.8% of cortical volume), we calculated tau pathology index (TPI):

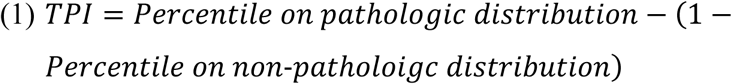

with the higher-mean distribution considered pathologic and the lower-mean distribution considered non-pathologic (or normal). This generates values between −100 and 100, where positive values indicate that a value lies further on the pathologic distribution than the non-pathologic distribution, while negative values indicate that a value lies more on the non-pathologic distribution than the pathologic distribution. The cutoff for Tau-positivity was set to 2 standard deviations (SDs) above the mean of the non-pathologic curve^23^. Three measures of tau were calculated for this study: 1) Mean Tau SUVR, 2) Tau Extent (% of cortex defined by all included ROIs that were Tau+), and 3) a combined measure of Tau Magnitude and eXtent (Tau-MaX, Figure 1):

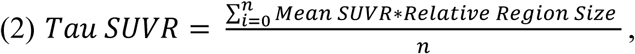

**Figure 1.**
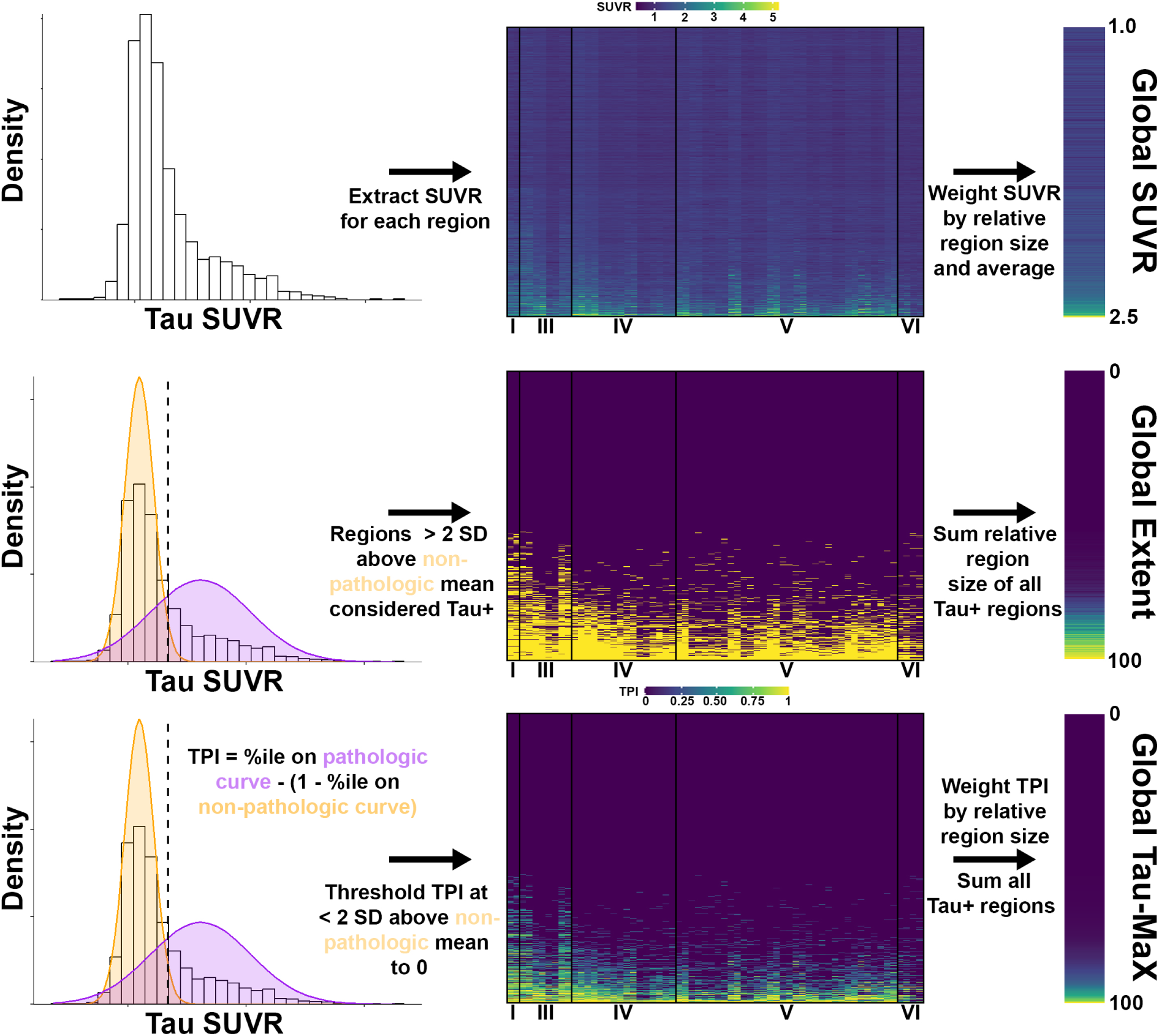
Measures of Tau Burden. Process for calculating global SUVR (*top*), Extent (*middle*), and Tau-MaX (*bottom*). The heatmap shows participants plotted along the y-axis and region organized by Braak stage on the x-axis. *Top*: Tau SUVR is extracted for each ROI and weighted by relative region size prior calculating a global average. *Middle*: Gaussian-mixture models (GMMs) are applied in each region to identify non-pathologic (orange) and pathologic (purple) distributions. A cutoff of 2 standard deviations (SD) above the mean of the non-pathologic distribution was used to define Tau+ regions. The sum of the relative region size of all Tau+ regions was calculated to generate an Extent measure ranging from 0 to 100 (percent). *Bottom*: The same GMMs as above are used to calculate Tau Pathology Index (TPI), which is then thresholded, such that all Tau regions are set to TPI = 0. TPI is weighted by relative region size and then summed across all Tau+ regions to calculate Tau-MaX, which ranges from 0 to 100.

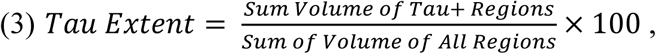

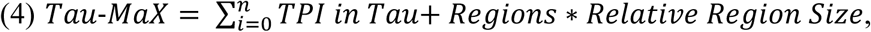

with relative regions size determined from region volume/volume of all included regions in the template brain. Each of these measures was calculated for four different sets of regions: 1) Global (all regions fit by the two-Gaussian model), 2) temporal meta-ROI ^13^, 3) Braak 1-4 meta-ROI^12^, and 4) Braak 5/6 meta-ROI^12^.

### Plasma Biomarker Analysis

A subset of participants had plasma p-tau_217_ values measured using Fujirebio Lumipulse® platform. Samples were collected and processed as previously described for ADNI^24^ and ABC^25^. All samples were analyzed on a Fujirebio Lumipulse G1200 analyzer at the University of Pennsylvania ADNI Biomarker Core laboratory ^24^.

### Neuropsychological Assessment

ADNI participants underwent standard neuropsychological assessment using MMSE and a battery of neuropsychological tests^26,27^. A measure of global cognition was calculated for ADNI participants by averaging the four composite scores available for memory, executive function, language, and visuospatial performance. The time point closest to Tau PET was used as baseline and the last available test was used for calculating of annualized change in global cognition. For ABC, all participants underwent neuropsychological assessment with the Uniform Dataset (UDS), which includes the MoCA^28–30^. A standard transformation was used to convert MoCA scores to derived MMSE scores^31^ for comparison to ADNI.

### Statistical Analyses

All statistical analyses were performed using R v4.4.1 (Packages: tidyverse, mixtools, dplyr, purr, ggplot2, scales, ggpubr, easystats, reshape, cocor, emmeans, lmerTest, knitr, kableExtra, vtable, broom, and fuzzySim).

### General Statistical Approach

Models using Global Tau-MaX, SUVR, and Extent were considered the primary outcome measure for all analyses, while analyses in other meta-ROIs were considered secondary/exploratory. Multiple comparison correction using Bonferroni or FDR adjusted *p-* values (specified by analysis in sections below) was performed within analyses but was not adjusted for repeating analyses in three additional meta-ROIs. Comparison between correlation coefficients used the Hittner’s Z dependent-group with overlapping variance model implemented in the *cocor* package^32^. Groups for analyses were defined by biological diagnosis incorporating clinical stage (CU, MCI, Dementia) and amyloid (Aβ) status. Analyses focused on the AD spectrum (CU Aβ-, CU Aβ+, MCI Aβ+, and AD). Two-tailed tests were used in all cases and adjusted *p* < 0.05 was considered significant.

### Factors Contributing to Cross-Sectional Tau Burden

We first assessed how tau magnitude and extent varied spatially across disease stages. T-tests were used to compare thresholded TPI (all Tau− regions set to TPI = 0) and extent (binary Tau− = 0 and Tau+ = 1) between sequential disease stages in the AD spectrum in each ROI separately. FDR-adjusted *p*-values were used to assess significance across the 64 regions tested. The average thresholded TPI and extent were calculated within each disease stage to further visualize alterations in magnitude and extent across the disease process. Additionally, individuals were divided into quintiles based on tau extent to visualize differences in magnitude across varying levels of spatial extent. Next the associations between SUVR, Extent, and Tau-MaX were explored within each disease stage separately using Pearson correlations. Hittner’s Z was used to compare correlation coefficients. Finally, we evaluated how each tau measure differed by Aβ status and clinical stage. Separate linear models including age and sex as covariates and the Aβ × Clinical Stage interaction as the predictor of interest were tested for each of the Tau measures as an outcome measure. Post-hoc t-tests were used to perform pairwise comparison between groups with Bonferroni correction for multiple comparisons.

### Longitudinal Change in Tau Burden

A subset of participants had multiple tau PET scans available, and linear mixed models were used to examine the longitudinal change in tau PET measures. For each global tau measure, we tested two models for longitudinal change across all participants while controlling for age and sex: 1) Time × Aβ status and 2) Time × Disease Stage. The primary analyses focused on global measures of tau and additional exploratory analyses were performed within each of the meta-ROIs (see Supplemental Materials). Unadjusted *p*-values were used to determine significance of interactions.

### Association with Plasma p-tau217, MMSE, and Global Neuropsychological Scores

Linear regression was used to assess the association of each tau measure with plasma p-tau_217_ in Aβ+ participants while controlling for age and sex. Similarly, linear regression was used to test the association of each tau measure with MMSE while controlling for age, sex, and education in MCI Aβ+ and AD participants. Hittner’s Z was used to compare correlation strengths within each ROI to identify the global tau measure with the strongest association with p-tau_217_ or MMSE. Analyses were repeated within meta-ROIs, and the numerically best measure in each meta-ROI was compared to the best global measure using Hittner’s Z.

Associations between global tau measure and global cognition were assessed in a subset of ADNI participants for whom cognitive measures were available. Association between global tau measures and global cognition were assessed cross-sectionally in Aβ+ participants using linear regression with the tau measure as the predictor of interest and age, sex, and education as covariates. These analyses were repeated using the annualized change in global cognition as the outcome measure, baseline tau measure as the predictor of interest, and age, sex, and education as covariates. Hittner’s Z was used to compare standardized regression coefficients of each global tau measure with cross-sectional and longitudinal cognition.

### Data availability

All requests for raw and analyzed data from the ABC cohort will be reviewed by the Penn Neurodegenerative Data Sharing Committee (PNDSC) and shared for appropriate uses through a data sharing agreement (https://www.pennbindlab.com/data-sharing). Anonymized data from ABC will be shared upon request to the corresponding author by a qualified academic investigator for the purpose of replicating procedures and results in this article. Data are not publicly available due to privacy protections outlined in the participant informed consent. Documents related to study protocols, informed consent and other documentation can similarly be made available upon request. All ADNI data are shared without embargo through the LONI Image and Data Archive (https://ida.loni.usc.edu/), a secure research data repository. Interested scientists may obtain access to ADNI imaging, clinical, genomic, and biomarker data for the purposes of scientific investigation, teaching, or planning clinical research studies. Access is contingent on adherence to the ADNI Data Use Agreement and the publications’ policies (https://adni.loni.usc.edu/data-samples/access-data/). All scripts for calculating Tau-MaX will be made available online upon publication and any additional code (imaging processing scripts, R scripts) used in these analyses is available upon request to the corresponding author.

## Results

### Participants

There were a total of 1077 participants with Tau PET data. At baseline, 606 participants were CU (177/600 Aβ+), 307 participants were MCI (157/304 Aβ+), 113 had Dementia (91/109 Aβ+), and 57 were impaired not MCI. The average time between Tau PET and Aβ PET was 63.4 ± 147.7 days. Full demographic data is available in Table 1. Plasma p-tau_217_ was available in 484 CU participants (145/483 Aβ+), 250 participants with MCI (130/237 Aβ+), and 92 participants with Dementia (74/88 Aβ+). The mean time between plasma p-tau_217_ and Tau PET was 349.6 ± 677.1 days. MMSE data (including converted MMSE) was available for 540 CU participants, 302 participants with MCI, and 113 participants with Dementia. The mean time between Tau PET and MMSE testing was 104.3 ± 315 days. Individuals who were missing age (n=4), amyloid status (n=13), or had diagnosis of impaired not MCI (n = 57) were included in GMMs for generation of Tau-MaX but were not included in additional analyses.

**Table 1.**
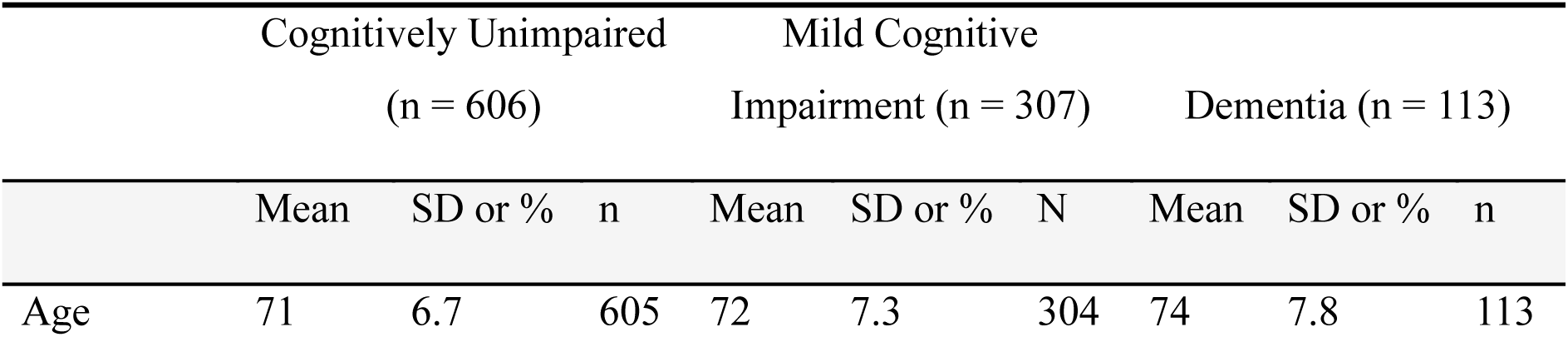
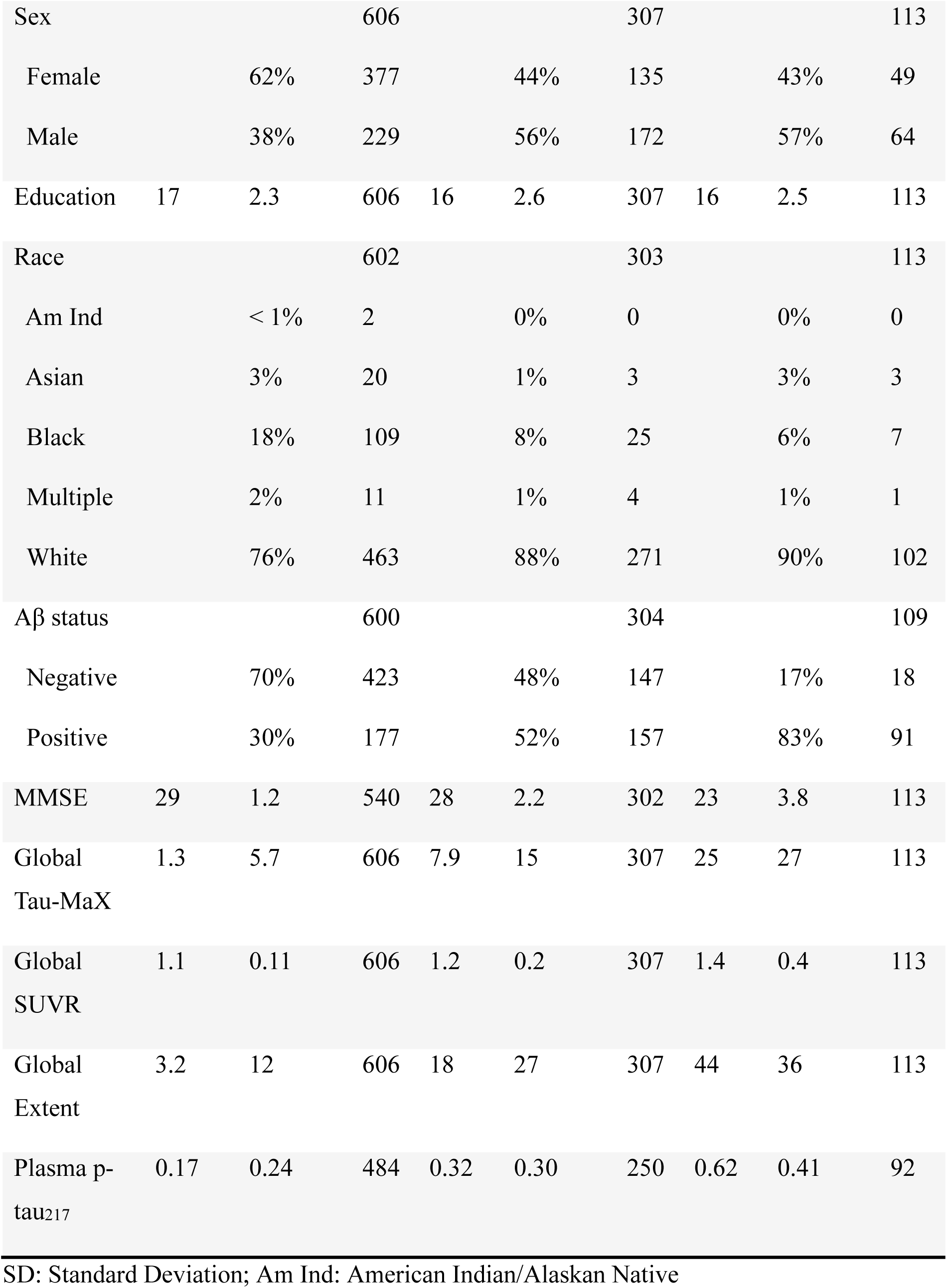
Demographics and summary measures.

### Tau Magnitude and Extent

Tau magnitude differed between clinical stages following a typical pattern of Braak staging with initial differences between CU Aβ+ and CU Aβ− confined to MTL and inferior temporal cortices, followed by greater increase in temporal cortices, angular gyrus, and posterior cingulate relative to frontal, parietal, and occipital increases in MCI Aβ+ compared to CU Aβ+ (Figure 2A). Differences in AD compared to MCI Aβ+ were less robust but showed similar patterns as between MCI Aβ+ and CU Aβ+. Tau extent largely followed similar pattern, but there were more pronounced effects at earlier stages with increases in temporal cortices and angular gyrus present in CU Aβ+ compared to CU Aβ−, as well as greater increases in posterior cingulate, frontal, and occipital cortices in MCI Aβ+ compared to CU Aβ+ (Figure 2A). Further differences in extent between AD and MCI Aβ+ were less pronounced than differences in magnitude. This pattern of earlier spread and later increases in magnitude was also present at the individual level when comparing SUVR across different quintiles of tau extent, with only small increases in magnitude until reaching the highest quintile of extent (Figure 2B-C).

**Figure 2.**
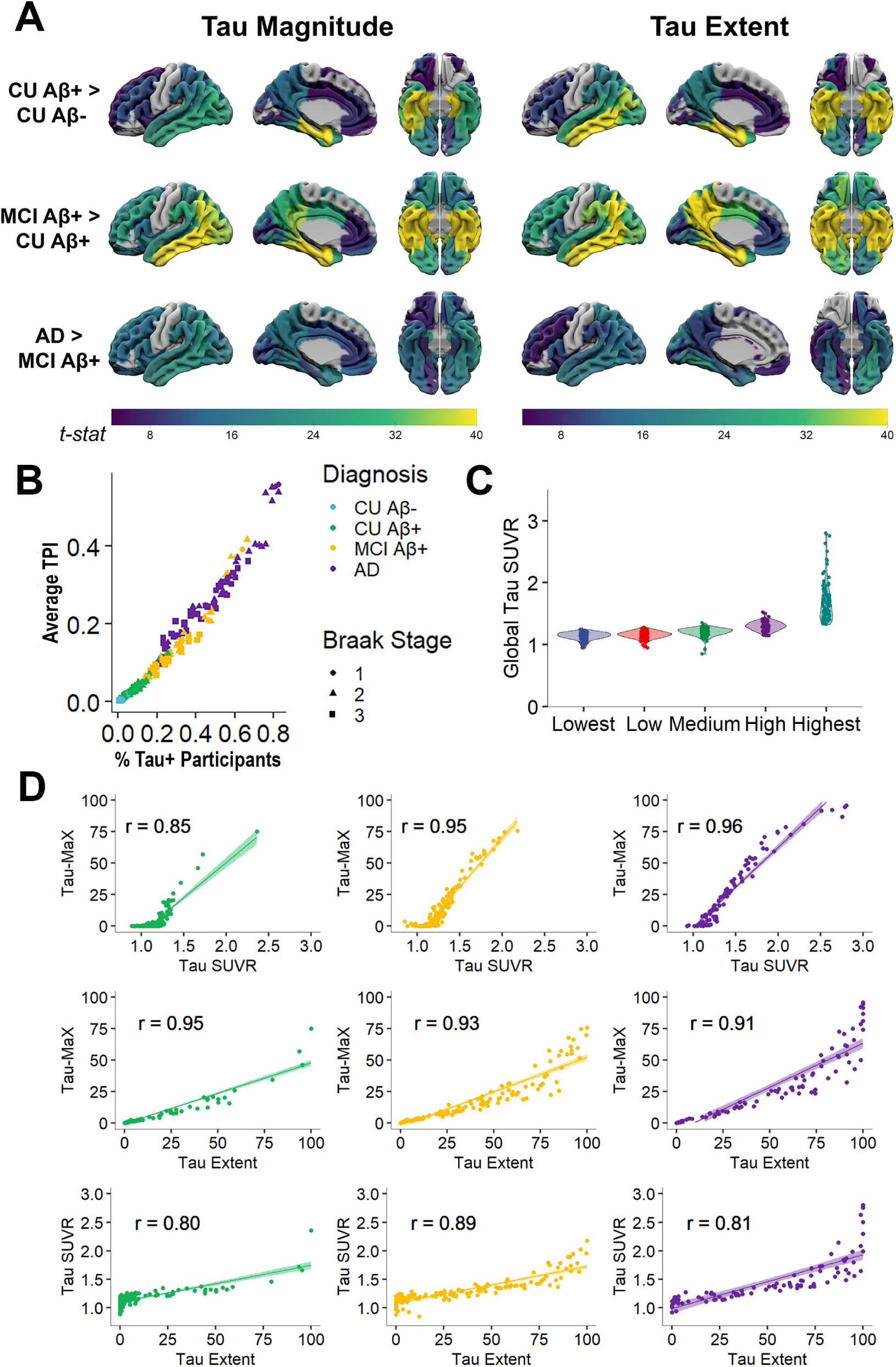
Tau Magnitude and Extent across the AD spectrum. **(A)** Tau magnitude (*left*) and extent (*right*) plotted on the MNI brain with right lateral, left medial, and inferior views shown. The color-scale reflects the *t*-statistic for the group comparison shown with the same scale used for all images shown. Only regions that were significant with FDR-corrected *p* < 0.05 are colored. (**B)** Regional averages plotted for each biological group (color scale) with shape depicting Braak stage (1 = Braak 1/2, 2 = Braak 3/4, 3 = Braak 5/6). The x-axis is the proportion of participants in the given biological group that were Tau+ in a region. The y-axis is the average TPI for a region in the given group after setting all Tau− regions to 0. **(C)** Global Tau SUVR is plotted with Tau+ participants (Tau Extent > 0) divided into groups based on quintile of tau extent. There is relatively low SUVR until the highest tau extent but there is high overlap between earlier groups and higher variability in the highest tau extent group. **(D)** Scatter plots of Tau-MaX with SUVR, (top), Tau-MaX with Extent (middle), and Extent with SUVR in CU Aβ+ (green), MCI (yellow), and AD (purple) separately. Pearson correlation coefficients are shown along with the linear best-fit (shaded region reflects the 95% confidence interval of fit).

We next evaluated if Tau-MaX was able to capture these dynamic changes in magnitude and extent across the disease spectrum in Aβ+ individuals (Figure 2D). Correlation analyses demonstrated a pattern of higher association between Tau-MaX and extent in CU (*r* = 0.95) that decreased in MCI (*r* = 0.93) and further in AD (*r* = 0.91), while the opposite pattern was seen for SUVR with lower association with Tau-MaX in CU (*r* = 0.85) that increased in MCI (*r* = 0.95) and AD (*r* = 0.96). This is also reflected by the weaker relationship between Extent and SUVR in CU (*r* = 0.80) and AD (*r* = 0.81) compared to MCI (*r* = 0.89). When comparing the strength of associations of Extent and Magnitude with Tau-MaX within each disease stage, Tau-MaX had a significantly stronger association with Extent compared to SUVR in CU (*Hittner’s Z* = −6.93, *P* < 0.001) and with SUVR compared to Extent in MCI (*Hittner’s Z* = 2.05, *P* = 0.04) and AD (*Hittner’s Z* = 4.10, *P* < 0.001).

### Factors Influencing Cross-Sectional Tau Burden

There was a significant Aβ status × Clinical stage interaction for all measures of global tau burden (*F*(2,1001) > 21.7, *P* < 0.001). Each measure showed the same pattern with higher tau burden in Aβ+ compared to Aβ− in all disease stages, no difference between tau burden across disease stages in Aβ− individuals, and significant differences between disease stages in Aβ+ individuals (Figure 3A). Specifically, Tau-MaX, SUVR, and Extent were higher in Aβ+ compared to Aβ− individuals in CU (*t*(1001) = 3.70, 4.54, 4.67, respectively, *P* < 0.001), MCI (*t*(1001) = 11.0, 9.61, 13.84, respectively, *P* < 0.001), and Dementia (*t*(1001) = 9.10, 7.29, 9.24, respectively, *P* < 0.001). Further Tau-MaX, SUVR, and Extent were higher in MCI than CU (*t*(1001) = 8.96, 7.09, 11.53, respectively, *P* < 0.001) and AD than MCI (*t*(1001) = 9.47, 8.03, 7.73, respectively, *P* < 0.001) in Aβ+ individuals. Similar findings were seen within the meta-ROIs (Supplemental Table 1).

**Figure 3.**
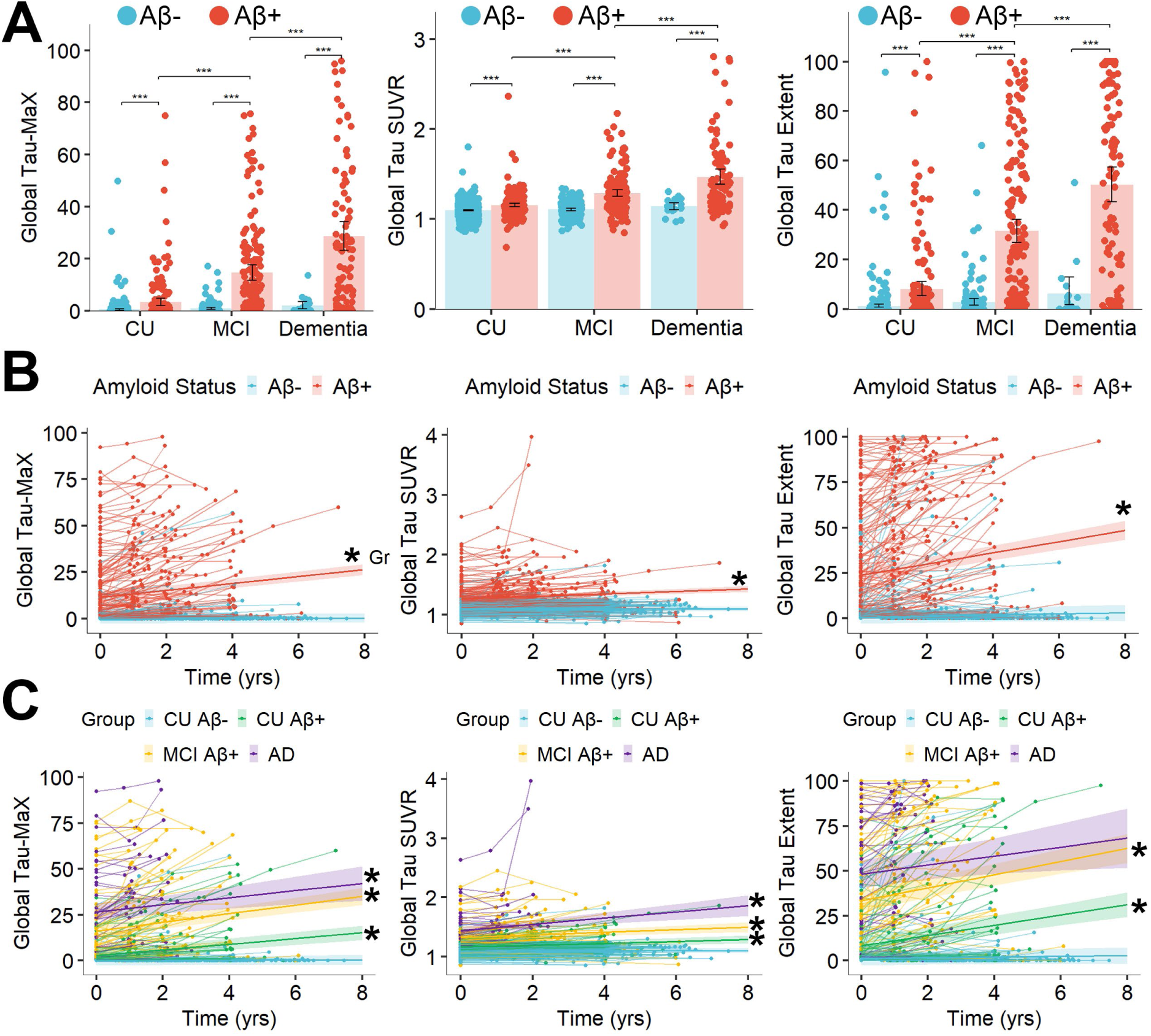
Measures of Global Tau along the AD spectrum. **(A)** Cross-sectional differences between diagnostic groups and amyloid status. Bars indicate the group mean with error bars representing the 95% confidence interval of mean. Significant differences are shown by brackets with \*\*\**P* < 0.001. **(B-C)** Spaghetti plots of longitudinal change by amyloid status **(B)** and diagnostic group **(C)** with thin lines connecting data points from the same participant and thick lines representing the linear best-fit for each group with shading reflecting 95% confidence interval of fit. \**P* < 0.05 for the Time × Group interaction. CU: Cognitively unimpaired, MCI: Mild Cognitive Impairment, AD: Alzheimer’s Disease.

### Longitudinal Change in Tau Burden

Longitudinal data was available for 453 individuals with an average 2.5 ± 0.8 scans per participant and average follow-up time of 2.6 ± 1.2 years. Results of linear-mixed models are shown in Figure 3B-C. Tau-MaX increased over time in Aβ+ individuals (*β* = 1.75 [1.40 – 2.10], *β_std_* = 0.18, *t*(1133) = 9.86, *P* < 0.001) but not Aβ− individuals (*P* = 0.27). When examining change over time by diagnostic group, Tau-MaX increased over time in CU Aβ+ (*β* = 1.46 [1.00 – 1.93], *β_std_* = 0.15, *t*(979) = 6.15, *P* < 0.001), MCI Aβ+ (*β* = 2.26 [1.72 – 2.79], *β_std_* = 0.23, *t*(979) = 8.24, *P* < 0.001), and AD (*β* = 1.83 [0.67 – 3.00], *β_std_* = 0.18, *t*(979) = 3.08, *P* = 0.002), but not in CU Aβ− (*P* = 0.43).

Similarly, Global SUVR increased over time in Aβ+ (*β* = 0.019 [0.013 – 0.026], *β_std_* = 0.14, *t*(1133) = 5.54, *P* < 0.001) but not in Aβ− individuals (*P* = 0.74). Further, SUVR increased over time in CU Aβ+ (*β* = 0.015 [0.006 – 0.24], *β_std_* = 0.10, *t*(979) = 3.24, *P* = 0.001), MCI Aβ+ (*β* = 0.021 [0.010 – 0.031], *β_std_* = 0.14, *t*(979) = 3.88, *P* < 0.001), and AD (*β* = 0.052 [0.029 – 0.075], *β_std_* = 0.36, *t*(979) = 4.44, *P* < 0.001), but not in CU Aβ− (*P* = 0.71). Global Extent also increased over time in Aβ+ (*β* = 2.80 [2.15 – 3.44], *β_std_* = 0.17, *t*(1133) = 8.56, *P* < 0.001) but not Aβ− individuals (*P* = 0.11). Finally, Extent increased over time in CU Aβ+ (*β* = 2.67 [1.83 – 3.51], *β_std_* = 0.16, *t*(979) = 6.24, *P* < 0.001), MCI Aβ+ (*β* = 3.45 [2.48 – 4.41], *β_std_* = 0.20, *t*(979) = 6.98, *P* < 0.001), and AD (*β* = 2.29 [0.19 – 4.39], *β_std_* = 0.13, *t*(979) = 4.39, *P* = 0.03) but not in CU Aβ− (*P* = 0.33).

### Association with Plasma p-Tau_217_

There was a total of 346 Aβ+ participants (defined by amyloid PET) with available plasma p-tau_217_. All measures of tau were significantly associated with plasma p-tau_217_ in Aβ+ individuals after controlling for age and sex (Figure 4). Plasma p-tau_217_ was strongly associated with Global Tau-MaX (*β_std_* = 0.70 [0.62 – 0.78], *P* < 0.001), SUVR (*β_std_* = 0.63 [0.54 – 0.72], *P* < 0.001), and Extent (*β_std_* = 0.68 [0.60 – 0.76], *P* < 0.001). Comparison of correlation strengths revealed that Tau-MaX had a significantly stronger association with p-tau_217_ than SUVR (*Hittner’s Z* = 5.91, *p* < 0.001) but only marginally stronger association than Extent (*Hittner’s Z* = 1.89, *p* = 0.059). Tau-MaX had the numerically strongest association with p-tau_217_ in all meta-ROIs, except temporal where Tau-MaX and SUVR were equivalent. The association of p-tau_217_ with Global Tau-MaX was significantly stronger than the association with Tau-MaX in temporal (*Hittner’s Z* = 2.05, *P* = 0.04), SUVR in temporal (*Hittner’s Z* = 1.99, *P* = 0.046), and Tau-MaX in Braak 1-4 (*Hittner’s Z* = 2.32, *P* = 0.02) meta-ROIs, but did not differ from the association with Tau-MaX in Braak 5/6 meta-ROI (*P* = 0.10).

**Figure 4.**
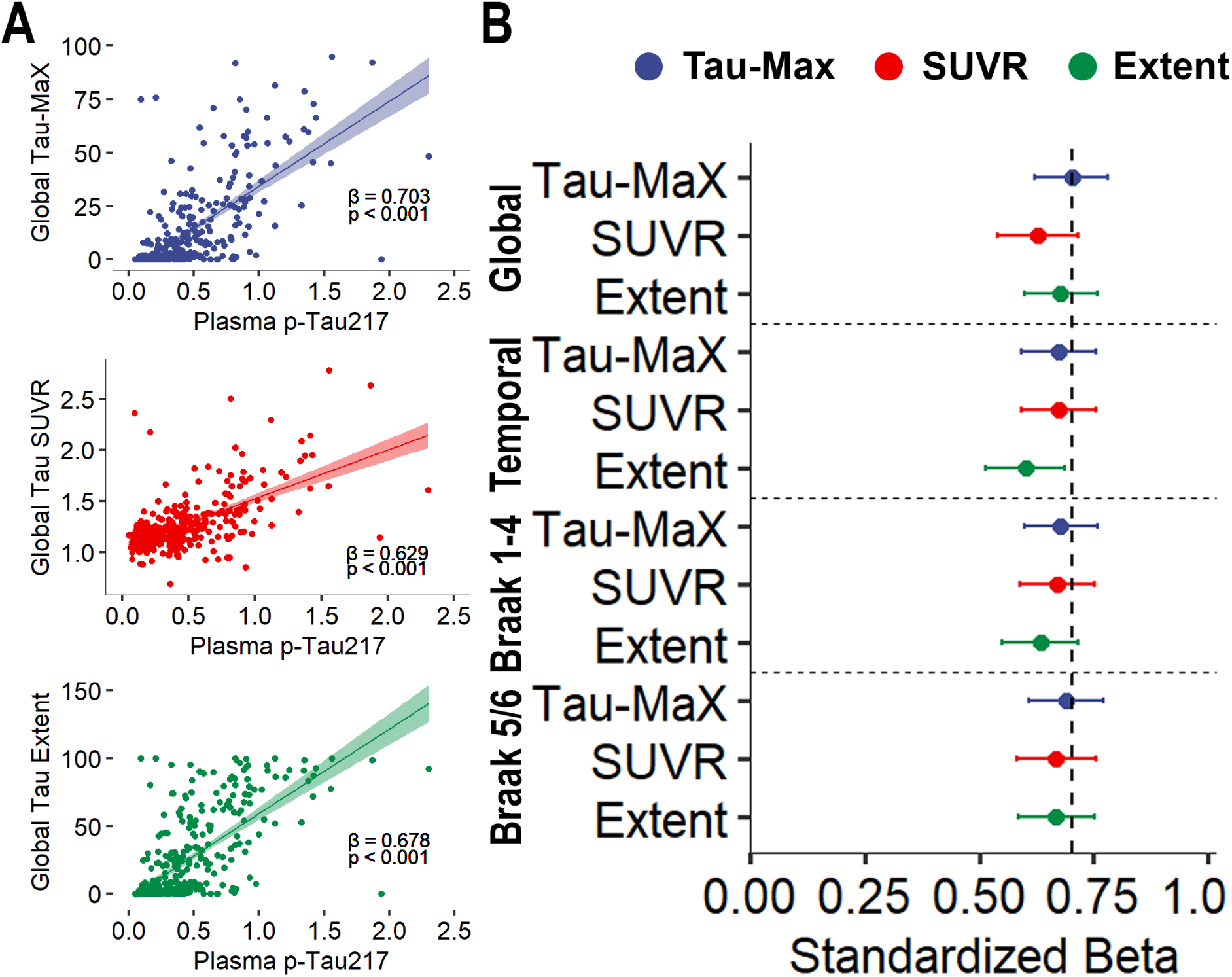
Association of tau burden with plasma p-tau_217_. **(A)** Associations between global measures of tau burden with plasma p-tau_217_ are shown with linear best-fit shown and shading representing 95% confidence of fit. Standardized β-values are shown. **(B)** Strength of standardized β-coefficients between plasma p-tau_217_ and measures of tau burden in meta-ROIs and globally. 95% confidence interval are shown. The dashed line is the relationship between Global Tau-MaX and plasma p-tau_217_.

### Association with MMSE

There were a total of 248 MCI Aβ+ and AD participants with available MMSE scores. All measures of tau were significantly associated with MMSE in MCI Aβ+ and AD individuals after controlling for age, sex, and education (Figure 5). MMSE was moderately negatively associated with Global Tau-MaX (*β_std_* = −0.49 [-0.61 – −0.37], *P* < 0.001), SUVR (*β_std_* = −0.46 [-0.58 – −0.33], *P* < 0.001), and Extent (*β_std_* = −0.44 [-0.56 – −0.32], *P* < 0.001). Comparison of correlation coefficients revealed that Tau-MaX had a significantly stronger association with MMSE than Global SUVR (*Hittner’s Z* = −1.99, *P* = 0.046) and Extent (*Hittner’s Z* = −2.31, *P* = 0.02). Tau-MaX had the numerically strongest association with MMSE in all meta-ROIs, except for the Temporal meta-ROI where SUVR had the numerically strongest association. The association of MMSE with Global Tau-MaX did not significantly differ from the association with SUVR in temporal (*P* = 0.21), Tau-MaX in Braak 1-4 (*P* = 0.24), or Braak 5/6 (*P* = 0.23) meta-ROIs.

**Figure 5.**
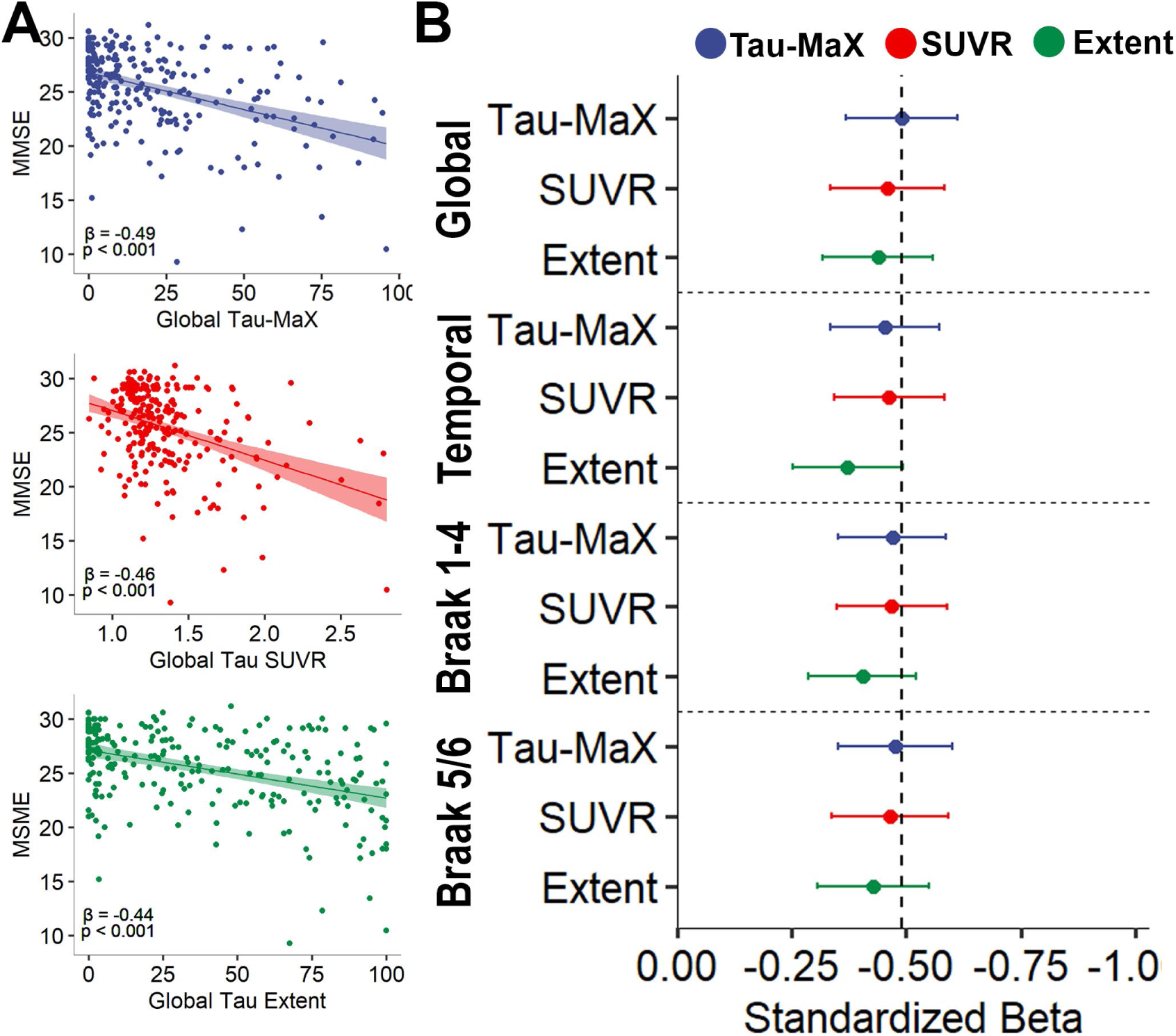
Association of tau burden with MMSE. **(A)** Associations between global measures of tau burden with MMSE are shown with linear best-fit shown and shading representing 95% confidence of fit. Standardized β-values are shown. **(B)** Strength of standardized β-values between MMSE and measures of tau burden in meta-ROIs and globally. 95% confidence intervals are shown. The dashed line is the relationship between Global Tau-MaX and MMSE.

### Association with Global Cognition on ADNI Neuropsychological Battery

A subset of 286 Aβ+ participants from ADNI had additional neuropsychological testing available. All measures of tau burden were significantly associated with the composite of global cognition after controlling for age, sex, and education (Figure 6A). Global cognition was strongly negatively associated with Global Tau-MaX (*β_std_* = −0.64 [-0.73 – −0.56], *t*(281) = −14.6, *P* < 0.001) and moderately negatively associated with SUVR (*β_std_* = −0.58 [-0.68 – - 0.49], *t*(281) = −12.3, *P* < 0.001) and Extent (*β_std_* = −0.59 [-0.68 – −0.50], *t*(281) = −13.1, *P* < 0.001). A follow-up linear-mixed model was run to assess for differences between Tau Measure and prediction of global cognition. Results demonstrated that there was a significant Measure × Cognition interaction such that Tau-MaX had a significantly stronger association with Global Cognition than SUVR (*t*(847) = 3.79, *P* < 0.001) and Extent (*t*(847) = 2.07, *P* = 0.038).

**Figure 6.**
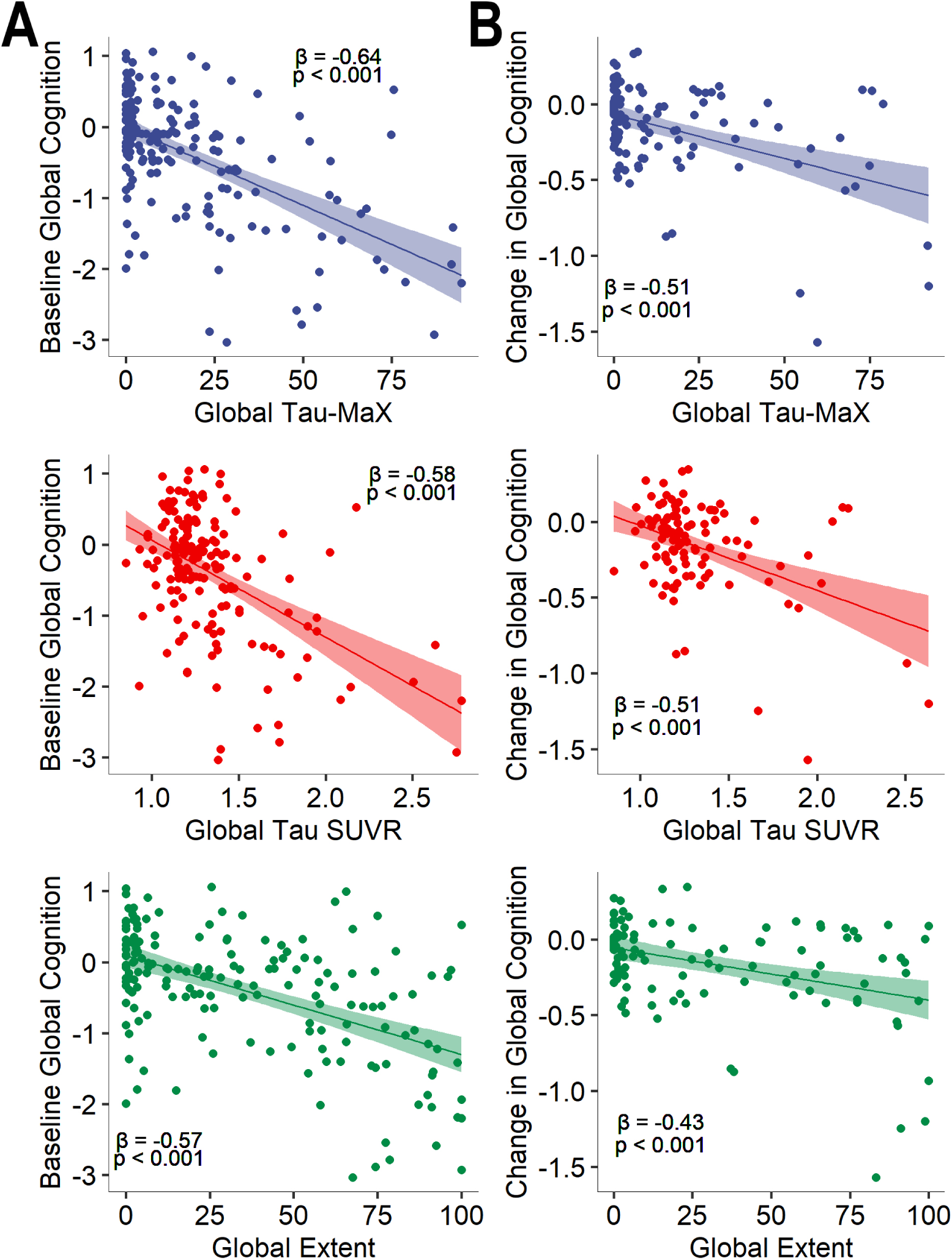
Association of Global Tau Burden with Global Cognition in ADNI. Associations between global measures of tau burden with global cognition at baseline **(A)** and with annualized change in global cognition **(B)** after controlling for age, sex, and education. Lines represent the linear best-fit with 95% confidence of fit shown by shading. Standardized beta-values are shown.

Longitudinal neuropsychological data was available for 184 Aβ+ participants. All measures of global tau were significantly associated with annualized change in global cognition after controlling for age, sex, and education (Figure 6B). Change in global cognition was moderately negatively associated with Global Tau-MaX (*β_std_* = −0.51 [-0.65 – −0.38], *t*(179) = −7.69, *P* < 0.001), SUVR (*β_std_* = −0.51 [-0.65 – −0.38], *t*(179) = −7.60, *P* < 0.001), and Extent (*β_std_* = −0.45 [-0.58 – −0.31], *t*(179) = −6.62, *P* < 0.001). A follow-up linear-mixed model was run to assess differences between Tau Measure and prediction of change in global cognition. Results demonstrated that there was a significant Measure × Cognition interaction, such that Tau-MaX had a significantly stronger association with change in Global Cognition than Extent (*t*(541) = 2.84, *P* = 0.005) but not SUVR (*t*(1.57), *P* = 0.12).

### Sensitivity Analysis

We replicated these analyses with comparable results using a cutoff of 2 SDs above the CU Aβ− mean SUVR as a threshold for Tau+ (see Supplemental Material).

## Discussion

We provide an extensive investigation of tau magnitude and extent across the disease spectrum, demonstrating earlier stages involve more prominent increases in number of Tau+ regions while later stages are dominated by accumulation in magnitude of tau burden in Tau+ regions. Combining these dimensions into a single measure of Tau Magnitude and eXtent (Tau-MaX) provides a robust metric of global tau burden that increases steadily throughout the disease process and shows strong associations with plasma p-tau_217_ and cognitive measures. Further, we demonstrate that global Tau-MaX outperformed or performed similarly to meta-ROI based measures of tau, thus allowing for region-agnostic assessment of tau burden. Together, these findings support the use of Tau-MaX as a robust measure of global tau burden.

Similar to prior tau PET and post-mortem studies, we found a stereotypical progression of both tau magnitude and extent closely resembling Braak stages with early changes in MTL regions followed by temporal neocortex and then later involvement of parietal, occipital, and frontal cortices^1,12,15^. We extend these prior studies by evaluating both magnitude and extent simultaneously, demonstrating earlier changes in extent followed by later increases in accumulation. Further, our finding of large variability in tau magnitude within a given level of extent, particularly at more advanced stages of spatial extent, are consistent with post-mortem findings of highly variable levels of tangle density within and across Braak stages^16–18^. Combining these two dimensions into a single measure provides a flexible metric of overall tau burden that both captures increase in extent in early stages of disease and accumulation of tau pathology within tau+ regions in later stages of disease.

Tau-MaX is sensitive to both cross-sectional differences along the AD spectrum, as well as longitudinal change in all stages of disease. SUVR and Extent also show the ability to detect cross-sectional differences and longitudinal change, although longitudinal change in SUVR was relatively weaker in CU Aβ+ and MCI Aβ+ compared to AD and change in Extent was only marginally significant in AD. A prior study using Spatial Extent Index did not find longitudinal change in extent in AD in a largely overlapping dataset, suggesting that our finding may be due to increased power from additional participants from ADNI and the ABC dataset rather than a stronger effect^15^. In addition, prior evaluations of global SUVR have similarly found greater longitudinal increases in later disease stages^33^. The flexibility of Tau-MaX appears robust to these differences with relatively stable changes longitudinally, likely capturing changes in extent in earlier stages and in SUVR in later stages.

Importantly, Global Tau-MaX was also associated with both a blood-based biomarker of AD pathology and cognition. Our findings are consistent with prior work demonstrating associations between p-tau_217_ and tau PET SUVR in medial temporal, temporoparietal, and late Braak stage regions^24,34,35^. We also extend prior findings of associations of plasma p-tau_217_ with Extent. Global Tau-MaX had the strongest association of all measures with plasma p-tau_217_ with only Global Extent and Braak 5/6 Tau-MaX showing non-inferior associations. To the degree that p-tau_217_ reflects global tau pathology, this result suggests Tau-MaX enhances measurement of overall tau burden compared to SUVR or Extent on their own. We also found associations between both cross-sectional and longitudinal cognition with Tau SUVR and Extent, consistent with prior studies^15,36–38^. Tau-MaX was more strongly associated with cross-sectional cognition than SUVR and with longitudinal change than Extent, again suggesting that the flexibility of Tau-MaX makes it adaptable to the relative strengths of each of these dimensions.

Overall, our findings support combining magnitude and extent into a single measure as a robust metric of global tau burden. There are several potential applications for such a metric. First, the revised criteria for AD diagnosis and staging use tau burden to delineate three of the four biological stages^11^. Tau-MaX provides a region-agnostic approach to staging tau severity, and future studies could evaluate potential cut-points that correspond to stages B-D in the staging system. Further, the continuous nature of Tau-MaX also allows for analyses that do not require binarization into these discrete categories. This may be of particular value for examining tau-clinical “mismatch” in which dissociations between clinical severity and tau burden may reflect resilience and vulnerability factors (e.g. presence of co-pathologies). Tau-MaX may be particularly useful for evaluating the effect of therapeutics on tau progression, as it does not require pre-specifying specific regions and can measure tau burden in heterogeneous populations. This may be beneficial for both clinical trials and studies of already approved disease modifying therapies.

In contrast, there may also be situations where combining these dimensions in a region-agnostic measure is not as valuable as examining these dimensions separately in specific regions of interest. Specifically, examining each of these dimensions as separate aspects of tau burden may be useful for identifying mechanisms related to tau spread versus accumulation. In addition, regional information is of particular importance for identifying disease subtypes and evaluating associations between tau and local neurodegeneration. There may also be important insights into different patterns of spreading and accumulation of tau pathology that differ on a regional basis that would be missed using a global, region-agnostic approach.

The current study has several limitations. First, it remains debated whether Gaussian mixture approaches or those using CU Aβ− individuals as the normal curve to develop a SUVR cutoff are preferred. Similar to prior studies of extent, we replicated our findings using CU Aβ− individuals to determine the Tau+ cutoff,^14^ suggesting that the method is robust to how the tau cutoff is determined. Next, the current study examines a single tracer, ^18^F-flortaucipir, using a single processing pipeline for combined data from ADNI and ABC. While the Tau-MaX approach should be applicable for other tracers and populations, future work is necessary to validate this measure in additional cohorts using multiple tau tracers. Further, our study primarily includes older adults who are CU or MCI with a smaller number of individuals with AD, and those with AD predominantly have late-onset AD. Additional studies in early-onset and atypical cases will be necessary to evaluate Tau-MaX in these settings, although it is expected that the region-agnostic approach of Tau-MaX will perform well in these cases. We also rely upon cognitive measures and blood-based biomarkers to assess the association with disease severity, but future studies comparing to post-mortem pathology will be valuable to determine the degree to which Tau-MaX may better capture the overall burden of tau pathology. In particular, comparing to quantitative neuropathology measures that assess both extent and density of pathology would be of high value to better understand how these dimensions are captured by *in vivo* PET imaging. Finally, examining Tau-MaX in populations from more diverse racial and ethnic backgrounds will be important to ensure generalizability of this approach to the wider population.

In conclusion, Tau-MaX provides a global measure of tau burden that captures dynamic changes in tau magnitude and extent across the disease spectrum. This allows for a region-agnostic approach to quantifying overall tau severity that is associated with blood-based biomarkers and cognition. Together, these findings provide strong support for Tau-MaX as an adaptive measure of global tau burden that may be of particular importance for disease monitoring in both natural history and in response to therapeutic interventions.

## Acknowledgements

Avid Radiopharmaceuticals, Inc., a wholly owned subsidiary of Eli Lilly and Company, enabled use of the 18F-flortaucipir tracer by providing precursor, but did not provide direct funding and was not involved in data analysis or interpretation

## Funding

This work was supported by grants from the National Institute of Health (P30-AG072979, RF1-AG069474, R01-AG056014, R01-AG055005, R01-AG072796, R25-NS065745, P01-AG084497), Pennsylvania Department of Health (2019NF4100087335), and Alzheimer’s Association and Fred A. and Barbara M. Erb Foundation (AACSF-23-1152241).

## Competing interests

CAB, SRD, JAD, PAY, KAQC, and CTM declare no competing interests. Ilya Nasrallah has served on the Scientific Advisory Board for Eisai and done educational speaking for Biogen. Alice Chen-Plotkin has a patent licensed to Prevail Therapeutics for genetic approaches to treating frontotemporal dementia. Leslie Shaw has served on scientific advisory boards and/or as a consultant for Biogen, Roche Diagnostics, Fujirebio, Siemens, and Diadem and has given lectures for Biogen, Roche, and Fujirebio. Edward Lee has served as a paid consultant for Wavebreak Therapeutics and Eli Lilly. David Wolk has served as a paid consultant for Eli Lilly and Beckman Coulter. He has also served on the DSMB for Functional Neuromodulation and GSK. He has received research support paid to his institution by Biogen.

Data collection and sharing for the Alzheimer’s Disease Neuroimaging Initiative (ADNI) is funded by the National Institute on Aging (National Institutes of Health Grant U19 AG024904). The grantee organization is the Northern California Institute for Research and Education. In the past, ADNI has also received funding from the National Institute of Biomedical Imaging and Bioengineering, the Canadian Institutes of Health Research, and private sector contributions through the Foundation for the National Institutes of Health (FNIH) including generous contributions from the following: AbbVie, Alzheimer’s Association; Alzheimer’s Drug Discovery Foundation; Araclon Biotech; BioClinica, Inc.; Biogen; Bristol-Myers Squibb Company; CereSpir, Inc.; Cogstate; Eisai Inc.; Elan Pharmaceuticals, Inc.; Eli Lilly and Company; EuroImmun; F. Hoffmann-La Roche Ltd and its affiliated company Genentech, Inc.; Fujirebio; GE Healthcare; IXICO Ltd.; Janssen Alzheimer Immunotherapy Research & Development, LLC.; Johnson & Johnson Pharmaceutical Research &Development LLC.; Lumosity; Lundbeck; Merck & Co., Inc.; Meso Scale Diagnostics, LLC.; NeuroRx Research; Neurotrack Technologies; Novartis Pharmaceuticals Corporation; Pfizer Inc.; Piramal Imaging; Servier; Takeda Pharmaceutical Company; and Transition Therapeutics.

## Supplementary material

Supplementary material is available online.

## Notes

### Funding Statement

This study was funded by grants from the National Institute of Health (P30-AG072979, RF1-AG069474, R01-AG056014, R01-AG055005, R01-AG072796, R25-NS065745, P01-AG084497), Pennsylvania Department of Health (2019NF4100087335), and Alzheimer's Association and Fred A. and Barbara M. Erb Foundation (AACSF-23-1152241) paid to the institutions of the authors.

### Author Declarations

IRB of University of Pennsylvania gave ethical approval for this work. Details about ADNI informed consent procedures are available from www.adni-info.org.

