## Supplemental Material for "Tau Burden is Best Captured by Magnitude and Extent: Tau-MaX as a Measure of Global Tau"

### Supplemental Results

#### Cross-Sectional Differences in Meta-ROI Tau Measures

There was a significant Aβ status × Clinical stage interaction for all measures of tau burden across all meta-ROIs ($F(2,1001) > 21.7$, $P < 0.001$). Each measure showed the same pattern with higher tau burden in Aβ+ compared to Aβ− in all disease stages, no difference between tau burden across disease stages in Aβ− individuals, and significant differences between disease stages in Aβ+ individuals (Table S1).

**Table S1. Meta-ROI Measures of Tau Burden**

| | | CU | | | MCI | | | Dementia | | | MCI Aβ+ > CU Aβ+ | AD > MCI Aβ+ |
|---|---|---|---|---|---|---|---|---|---|---|---|---|
| | | Aβ− | Aβ+ | Aβ+ > Aβ− | Aβ− | Aβ+ | Aβ+ > Aβ− | Aβ− | Aβ+ | Aβ+ > Aβ− | | |
| Temp-oral | Tau-MaX | 0.11<br>(0.73) | 6.03<br>(1.13) | 4.47<br>(< 0.001) | 1.17<br>(1.23) | 25.1<br>(1.20) | 13.9<br>(< 0.001) | 4.03<br>(3.50) | 42.5<br>(1.57) | 10.1<br>(< 0.001) | 11.6<br>(< 0.001) | 8.88<br>(< 0.001) |
| | SUVR | 1.17<br>(0.013) | 1.29<br>(0.021) | 4.93<br>(< 0.001) | 1.20<br>(0.022) | 1.59<br>(0.022) | 12.4<br>(< 0.001) | 1.29<br>(0.064) | 1.90<br>(0.029) | 8.68<br>(< 0.001) | 9.83<br>(< 0.001) | 8.63<br>(< 0.001) |
| | Extent | 1.41<br>(1.15) | 14.2<br>(1.78) | 6.07<br>(< 0.001) | 4.33<br>(1.93) | 49<br>(1.88) | 16.7<br>(< 0.001) | 9.05<br>(5.49) | 71<br>(2.46) | 10.4<br>(< 0.001) | 13.6<br>(< 0.001) | 7.11<br>(< 0.001) |
| Braak 1-4 | Tau-MaX | 0.07<br>(0.67) | 5.24<br>(1.03) | 4.21<br>(< 0.001) | 1.20<br>(1.12) | 21.5<br>(1.09) | 12.9<br>(< 0.001) | 4.16<br>(3.19) | 37.6<br>(1.43) | 9.58<br>(< 0.001) | 10.8<br>(< 0.001) | 9.02<br>(< 0.001) |
| | SUVR | 1.14<br>(0.012) | 1.24<br>(0.018) | 4.82<br>(< 0.001) | 1.16<br>(0.019) | 1.48<br>(0.019) | 11.7<br>(< 0.001) | 1.24<br>(0.055) | 1.75<br>(0.025) | 8.47<br>(< 0.001) | 9.22<br>(< 0.001) | 8.72<br>(< 0.001) |
| | Extent | 1.40<br>(1.15) | 12.3<br>(1.78) | 5.60<br>(< 0.001) | 4.49<br>(1.93) | 43.3<br>(1.88) | 15.7<br>(< 0.001) | 9.94<br>(5.49) | 64.2<br>(2.46) | 9.82<br>(< 0.001) | 13.1<br>(< 0.001) | 7.37<br>(< 0.001) |
| Braak 5/6 | Tau-MaX | 0.00<br>(0.57) | 3.44<br>(0.87) | 3.32<br>(< 0.001) | 0.45<br>(0.95) | 13.1<br>(0.92) | 9.56<br>(< 0.001) | 1.97<br>(2.71) | 27.0<br>(1.21) | 8.47<br>(< 0.001) | 7.65<br>(< 0.001) | 9.15<br>(< 0.001) |
| | SUVR | 1.09<br>(0.011) | 1.17<br>(0.017) | 4.26<br>(< 0.001) | 1.10<br>(0.019) | 1.34<br>(0.018) | 9.01<br>(< 0.001) | 1.17<br>(0.053) | 1.60<br>(0.024) | 7.56<br>(< 0.001) | 6.58<br>(< 0.001) | 9.00<br>(< 0.001) |
| | Extent | 1.35<br>(1.00) | 8.79<br>(5.75) | 5.77<br>(< 0.001) | 3.33<br>(1.68) | 31.7<br>(1.65) | 12.1<br>(< 0.001) | 6.61<br>(4.78) | 49.3<br>(2.14) | 8.15<br>(< 0.001) | 10.2<br>(< 0.001) | 6.53<br>(< 0.001) |

Group estimated marginal mean (standard error), *t*-statistic (*P*-value) for group comparisons; CU: cognitively unimpaired, MCI: Mild Cognitive Impairment

#### Longitudinal Change in Tau

Patterns of longitudinal change in meta-ROIs showed similar trends is global measures except for slight variance in patterns seen in earlier meta-ROIs (temporal, Braak 1-4) compared to the Braak

5/6 meta-ROI (Table S2). There were longitudinal increases in all measures of tau in all meta-ROIs in Aβ+ individuals but increases in Aβ- individuals were only seen in temporal and Braak 1-4 Extent. Similarly, there were no longitudinal increases in any tau measure in any meta-ROI in CU Aβ- individuals, but there were longitudinal increases in CU Aβ+, MCI Aβ+, and AD in all regions except for extent in Braak 5/6 in AD. There was a pattern of maximal longitudinal increases in MCI for all measures in temporal and Braak 1-4 meta-ROIs with smaller change in individuals with AD. In contrast, Tau-MaX and Extent increases in Braak 5/6 meta-ROI peaked in MCI, while SUVR increases in Braak 5/6 peaked in AD.

**Table S2. Longitudinal change in Tau in Meta-ROIs**

| | | Aβ− | Aβ+ | CU Aβ− | CU Aβ+ | MCI Aβ+ | AD |
|---|---|---|---|---|---|---|---|
| Temp-oral | Tau-MaX | 0.17 [-0.07, 0.42] | 2.34 [1.92, 2.75] | 0.12 [-0.16, 0.41] | 2.03 [1.48, 2.59] | 3.08 [2.44, 3.71] | 1.73 [0.34, 3.11] |
| | | $t$ = 1.41 (0.16) | $t$ = 11.0 (<0.001) | $t$ = 0.86 (0.39) | $t$ = 7.20 (<0.001) | $t$ = 9.46 (<0.001) | $t$ = 2.45 (0.014) |
| | SUVR | 0.004 [-0.000, 0.008] | 0.04 [0.03, 0.04] | 0.004 [-0.000, 0.009] | 0.03 [0.02, 0.04] | 0.05 [0.04, 0.06] | 0.03 [0.01, 0.06] |
| | | $t$ = 1.81 (0.07) | $t$ = 9.72 (< 0.001) | $t$ = 1.65 (0.098) | $t$ = 5.96 (< 0.001) | $t$ = 8.14 (<0.001) | $t$ = 2.48 (0.013) |
| | Extent | 0.47 [0.06, 0.89] | 2.52 [1.80, 3.24] | 0.28 [-0.17, 0.74] | 2.79 [1.89, 3.69] | 2.95 [1.92, 3.98] | 2.54 [0.30, 4.78] |
| | | $t$ = 2.23 (0.026) | $t$ = 6.89 (< 0.001) | $t$ = 1.21 (0.23) | $t$ = 6.10 (< 0.001) | $t$ = 5.60 (< 0.001) | $t$ = 2.23 (0.026) |
| Braak 1-4 | Tau-MaX | 0.16 [-0.07, 0.39] | 2.25 [1.84, 2.65] | 0.12 [-0.15, 0.39] | 1.88 [1.35, 2.41] | 2.98 [2.37, 3.60] | 2.12 [0.78, 3.45] |
| | | $t$ = 1.34 (0.18) | $t$ = 10.9 (<0.001) | $t$ = 0.85 (0.40) | $t$ = 6.91 (<0.001) | $t$ = 9.52 (<0.001) | $t$ = 3.12 (0.002) |
| | SUVR | 0.003 [-0.000, 0.007] | 0.032 [0.026, 0.039] | 0.003 [-0.001, 0.008] | 0.03 [0.02, 0.04] | 0.04 [0.03, 0.05] | 0.03 [0.01, 0.06] |
| | | $t$ = 1.63 (0.10) | $t$ = 9.44 (<0.001) | $t$ = 1.46 (0.15) | $t$ = 5.72 (<0.001) | $t$ = 7.90 (<0.001) | $t$ = 2.90 (0.004) |
| | Extent | 0.41 [0.2, 0.79] | 2.88 [2.22, 3.54] | 0.24 [-0.18, 0.66] | 2.75 [1.92, 3.57] | 3.60 [2.65, 4.56] | 3.15 [1.09, 5.21] |
| | | $t$ = 2.09 (0.037) | $t$ = 8.55 (<0.001) | $t$ = 1.13 (0.26) | $t$ = 6.53 (<0.001) | $t$ = 7.44 (<0.001) | $t$ = 3.00 (0.002) |
| Braak 5/6 | Tau-MaX | 0.08[-0.15, 0.30] | 1.49 [1.11, 1.88] | 0.07 [-0.19, 0.34] | 1.30 [0.78, 1.82] | 1.82 [1.21, 2.42] | 1.77 [0.46, 3.07] |
| | | $t$ = 0.68 (0.50) | $t$ = 7.60 (<0.001) | $t$ = 0.54 (0.59) | $t$ = 4.87 (<0.001) | $t$ = 5.92 (<0.001) | $t$ = 2.66 (0.008) |
| | SUVR | 0.000 [-0.004, 0.006] | 0.02 [0.01, 0.03] | 0.001 [-0.005, 0.007] | 0.02 [0.004, 0.03] | 0.02 [0.01, 0.04] | 0.07 [0.04, 0.10] |
| | | $t$ = 0.23 (0.82) | $t$ = 4.95 (< 0.001) | $t$ = 0.17 (0.86) | $t$ = 2.74 (0.006) | $t$ = 3.46 (0.001) | $t$ = 4.70 (<0.001) |
| | Extent | 0.21 [-0.23, 0.65] | 2.83 [2.07, 3.59] | 0.17 [-0.34, 0.69] | 2.75 [1.74, 3.76] | 3.36 [2.20, 4.52] | 2.11 [-0.41, 4.64] |
| | | $t$ = 0.94 (0.35) | $t$ = 7.34 (< 0.001) | $t$ = 0.66 (0.51) | $t$ = 5.35 (< 0.001) | $t$ = 5.67 (< 0.001) | $t$ = 1.64 (0.10) |

Unstandardized β-values reflecting change over time are shown with [95% CI], *t*-statistic (*P*-value)

#### Alternative Tau+ Cutoff Based on CU Aβ− participants

Primary analyses were repeated using a cutoff for Tau+ of 2 SD's above the mean of the CU Aβ− participants. Similar results were seen as when using the GMM cutoff. Each measure showed the same pattern with higher tau burden in Aβ+ compared to Aβ− in all disease stages, no difference between tau burden across disease stages in Aβ− individuals and significant differences between stages in Aβ+ individuals (Figure S1A). The same pattern of longitudinal increase was seen as when using the GMM cutoff: all measure of global tau burden increased over time in Aβ+ but not

Aβ− participants and across all Aβ+ disease stages aside from Extent in individuals with Dementia (Figure S1B-C).

**Figure S1**

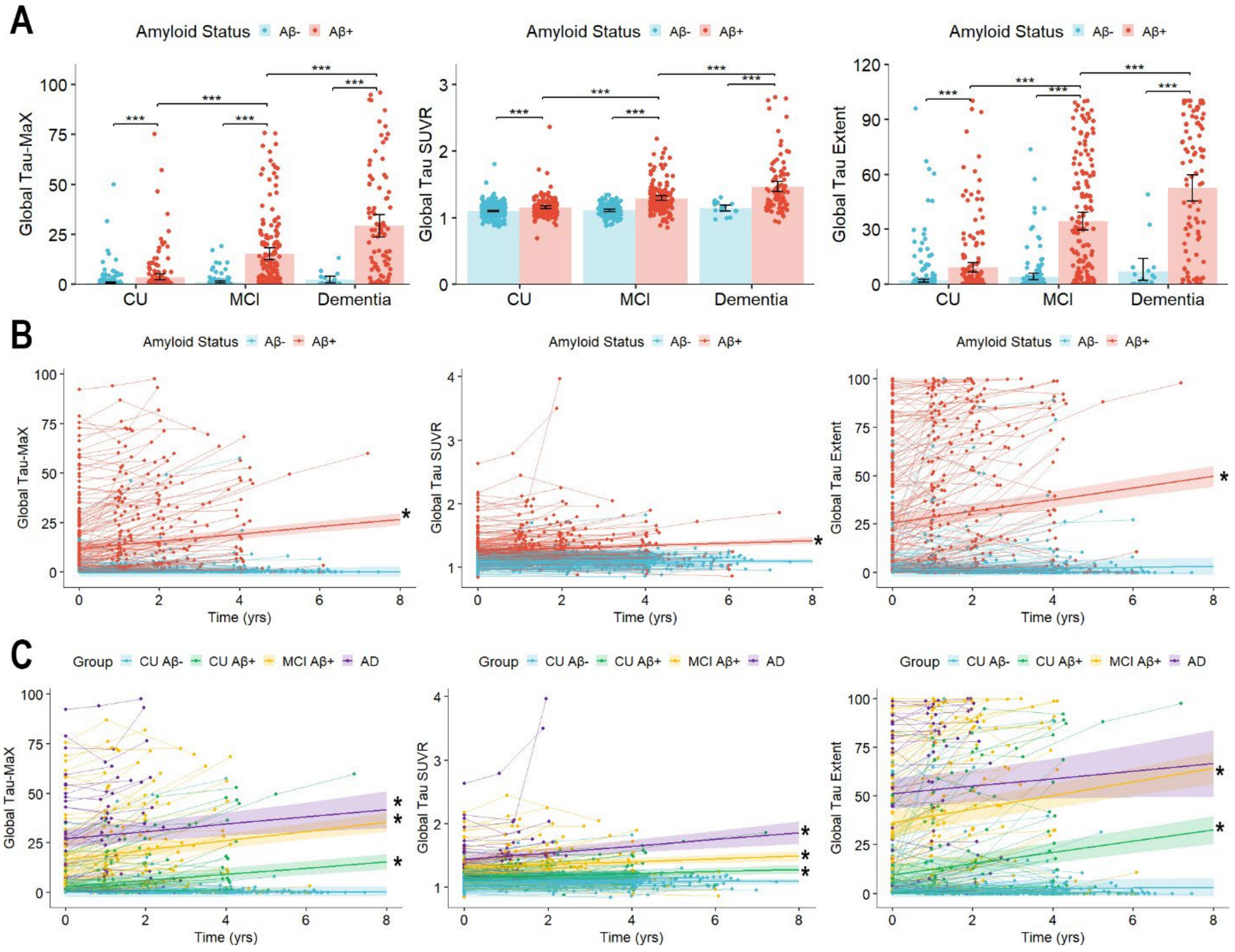


**Figure S1. Measures of Global Tau along the AD spectrum using CU AB- cutoff. (A)** Cross-sectional differences between diagnostic groups and amyloid status. Bars indicate the group mean with error bars representing the 95% confidence interval of mean. Significant differences are shown by brackets with ***$P < 0.001$. **(B-C)** Spaghetti plots of longitudinal change by amyloid status **(B)** and diagnostic group **(C)** with thin lines connecting data points from the same participant and thick lines representing the linear best-fit for each group with shading reflecting 95% confidence interval of fit. *$P < 0.05$. CU: Cognitively unimpaired, MCI: Mild Cognitive Impairment, AD: Alzheimer's Disease.

There was a significant positive association between all measures of tau burden and plasma p-tau$_{217}$ in Aβ+ participants, as well as a significant negative association between all measures of tau burden and MMSE in cognitively impaired Aβ+ participants (Figure S2). Global Tau-MaX had the strongest association with p-tau$_{217}$ (*Hittner's* $Z > 2.46$, $P \leq 0.014$) compared to SUVR and Extent. It had the numerically strongest association with MMSE as well but was not significantly stronger than SUVR or Extent (*Hittner's* $Z \geq 1.62$, $P \leq 0.10$). In addition, Tau-MaX had the numerically highest association with p-tau$_{217}$ and MMSE in all meta-ROIs except for Temporal meta-ROI, where SUVR was numerically higher than Tau-MaX.

**Figure S2**

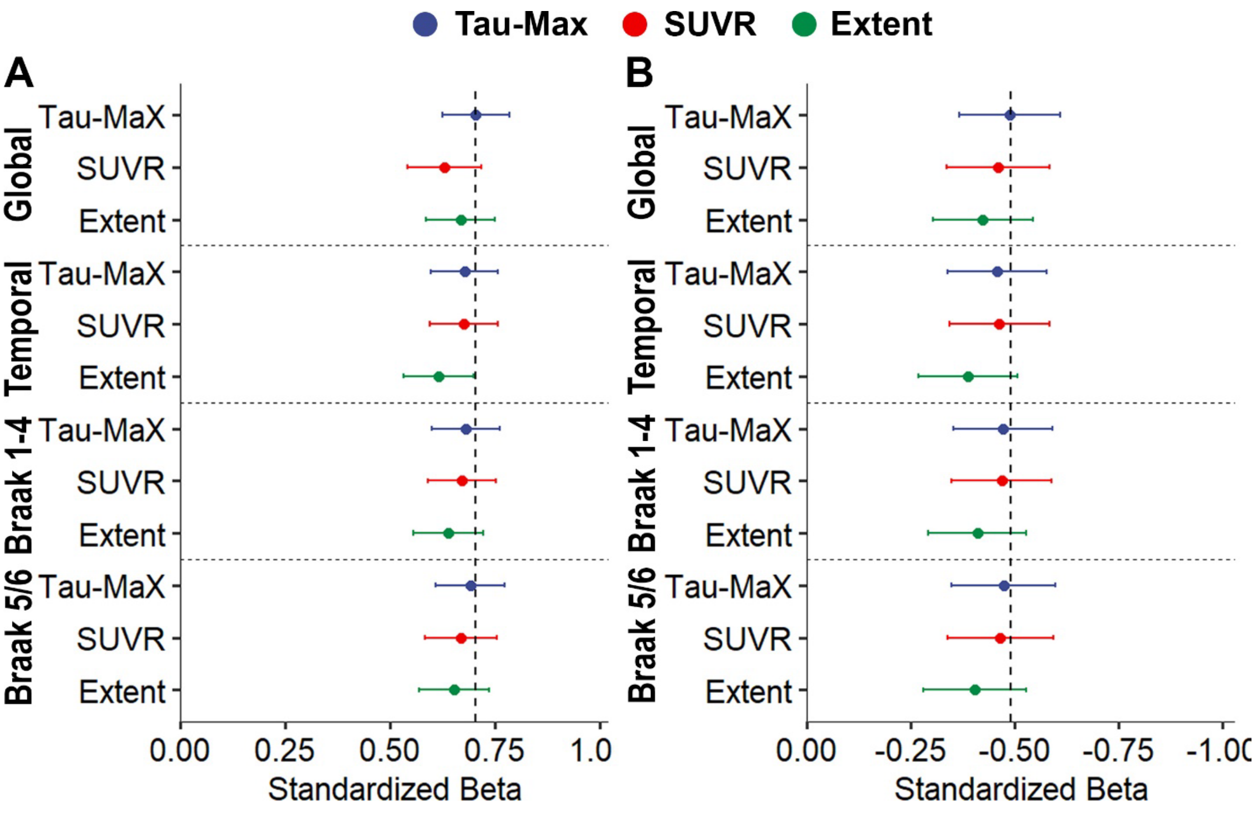


**Figure S2. Association of tau burden with plasma p-tau$_{217}$ and MMSE using CU Aβ- cutoff**. Strength of standardized β-coefficient of plasma p-tau$_{217}$ **(A)** and MMSE **(B)** with measures of tau burden in meta-ROIs and globally. 95% confidence interval of spearman coefficient are shown. The dashed line is the relationship between Global Tau-MaX and plasma p-tau$_{217}$ **(A)** or MMSE **(B)**.

Finally, Global Tau-MaX was a strong predictor of cross-sectional global cognition ($\beta_{std}$ = -0.64 [-0.73 – -0.56], $t(281) = -14.6$, $P < 0.001$) and a moderate predictor of longitudinal change in global cognition ($\beta_{std}$ = -0.51 [-0.64 – -0.38], $t(281) = -7.61$, $P < 0.001$). There was a significant Measure × Cognition interaction, such that there was a stronger association between baseline cognition and Tau-MaX compared to SUVR ($t(847) = 3.59$, $P < 0.001$) and Extent ($t(847) = 2.61$, $P = 0.009$). For longitudinal cognition, there was again a Measure × Cognition interaction such that there was a stronger association between longitudinal change in global cognition and Tau-MaX compared to Extent ($t(541) = 3.28$, $P = 0.001$) but not SUVR ($t(541) = 1.34$, $P = 0.18$). Relationships of each measure with global cognition are shown in Figure S3.

**Figure S3.**

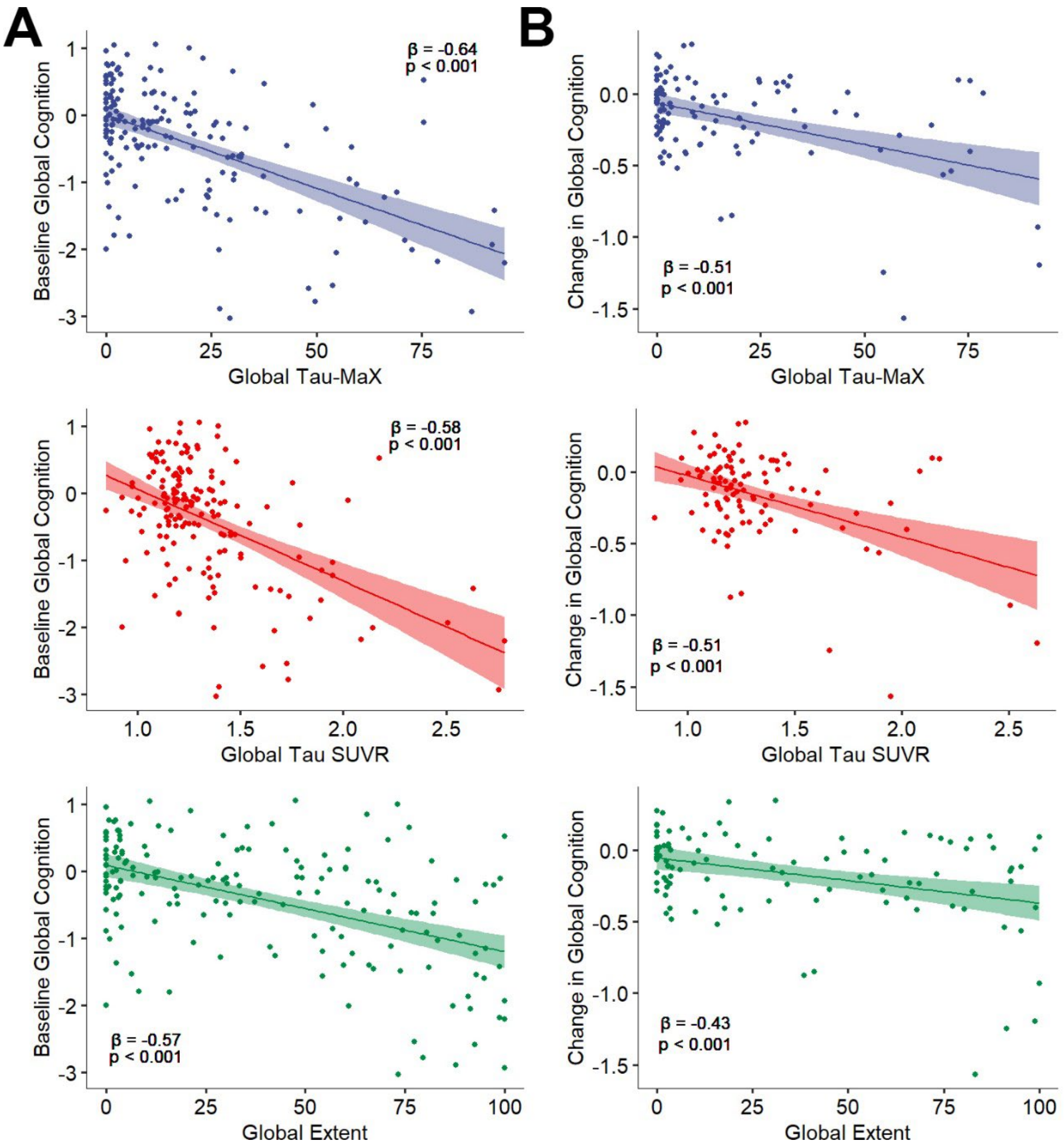


**Figure S3. Association of Global Tau Burden with Global Cognition in ADNI using Aβ-cutoff.** Associations between global measures of tau burden with global cognition at baseline (A) and with annualized change in global cognition (B) after controlling for age, sex, and education. Lines represent the linear best-fit with 95% confidence of fit shown by shading. Standardized beta-values are shown.
